# Disentangling the common genetic architecture and causality of rheumatoid arthritis and systemic lupus erythematosus with COVID-19 outcomes: genome-wide cross trait analysis and bi-directional Mendelian randomization study

**DOI:** 10.1101/2022.06.10.22276268

**Authors:** Minhao Yao, Xin Huang, Yunshan Guo, Jie V. Zhao, Zhonghua Liu

## Abstract

COVID-19 may cause a dysregulation of the immune system and has complex relationships with multiple autoimmune diseases, including rheumatoid arthritis (RA) and systemic lupus erythematosus (SLE). However, little is known about their common genetic architecture. We analysed summary-level genetic data from the latest COVID-19 host genetics consortium and consortia on RA and SLE to examine the shared genetic etiology and causal relationship between COVID-19 and RA/SLE. The cross-trait meta-analysis identified 46, 47, and 19 shared genetic loci for severe COVID-19, COVID-19 hospitalization, and SARS-CoV-2 infection with RA, and 19, 24, and 11 shared loci with SLE, respectively. Shared genes were significantly enriched in the spleen, lung, whole blood, and small intestine, and involved in immune function, inflammation and coagulation process. Co-localization analysis identified eight shared loci in *TYK2, IKZF3, COL11A2, PSORS1C1, MANEAL* and *COG6* genes for COVID-19 with RA, and four in *CRHR1, FUT2* and *NXPE3* genes for COVID-19 with SLE. Bi-directional Mendelian randomization analysis suggested RA is associated with a higher risk of COVID-19 hospitalization, and COVID-19 is not related to RA or SLE. Our novel findings improved the understanding of the common genetic aetiology shared by COVID-19, RA and SLE, and suggested an increased risk of COVID-19 hospitalization in people with higher genetic liability to RA.

## Introduction

Coronavirus Disease (COVID-19) pandemic poses a heavy burden on global health. Immune hyperactivation and cytokine storm have been identified as the key contributors to severe COVID-19. Meanwhile, immune dysfunction also leads to autoimmune diseases, including rheumatoid arthritis (RA) and systemic lupus erythematosus (SLE). Therefore, RA and SLE may have common genetic architecture with COVID-19. Identifying their shared genetic architecture can provide more insight into the complex molecular mechanisms underlying COVID-19 and auto-immune diseases. Although large-scale genome-wide association studies (GWASs) have been performed to understand the genetic architecture of RA, SLE and COVID-19 related traits (severe COVID-19, COVID-19 hospitalization, and SARS-COV-2 infection), to our knowledge, their shared genetic architecture has not been investigated.

Moreover, the causal relationship between COVID-19 and RA/SLE has not been clarified. In a large observational study, OpenSAFELY, in 17,278,392 adults with 10,926 COVID-19-related deaths, autoimmune diseases are related to a higher risk of COVID-19 related deaths.^1^ Current evidence from epidemiological studies is inconsistent and limited. For example, although autoimmune diseases are associated with a higher risk of severe COVID-19,^1^ there are also studies showing similar risk of hospitalization and mortality in people with or without RA,^2^ and no association of SLE with the risk of contracting COVID-19.^3^ On the other hand, there are concerns that COVID-19 may trigger autoimmunity and thereby induce RA or SLE, whether the occurrence of RA or SLE after COVID-19 is coincidence or causally associated has not been determined due to the limited number of cases and follow-up time.^4,5^ Observational studies are also subject to unmeasured confounding bias,^6^ which limits drawing causal conclusions. Given the obscure evidence, provisional guidelines from European League Against Rheumatism (EULAR), American College of Rheumatology (ACR) and those from Germany and UK, as well as clinical relevant studies, concluded that there was no evidence that patients with RA or SLE should be managed differently,^7,8^ but the “blind flying” calls for more studies in this research area for evidence-based clinical practice. To resolve this issue, Mendelian randomization (MR) study uses genetic variants as instrument for causal inference and can provide unconfounded estimates in an observational setting even in the presence of unmeasured confounding.^9^

To fill this urgent gap, we conducted a large-scale genome-wide cross-trait analysis to estimate their genetic correlations and shared genetic components between COVID-19 related traits and RA or SLE using publicly available summary level data. We further used two-sample bi-directional Mendelian randomization methods to assess the causal associations between COVID-19 related traits and RA or SLE.

## Results

### Genetic correlation of RA and SLE with COVID-19 related traits

The heritability (h^2^) estimated by LDSC suggested severe COVID-19, COVID-19 hospitalization, SARS-CoV-2 infection, RA and SLE are heritable (P<0.05 as shown in Supplementary Table 1). We found significant positive genetic correlations of COVID-19 hospitalization and SARS-CoV-2 infection with RA (r_g_ = 0.1252, P-value = 0.0084 for COVID-19 hospitalization with RA; r_g_ = 0.1552, P-value= 0.0088 for SARS-CoV-2 infection with RA). The positive genetic correlations of severe COVID-19 with RA and SLE (r_g_ =0.0938 for severe COVID-19 with RA; r_g_ = 0.0873 for severe COVID-19 with SLE), COVID-19 hospitalization with SLE (r_g_ = 0.0665), and SARS-CoV-2 infection with SLE (r_g_ = 0.0110) were not significant (P>0.05) (Figure 1 and Supplementary Table 2). Further partitioned LDSC analysis found that COVID-19 hospitalization is significantly associated with RA in the conserved region, DGF, DHS, H3K9ac, H3K27ac and transcribed region, and that severe COVID-19 and SARs-CoV-2 infection are associated with RA in the conserved region and transcribed region, respectively (Figure 2A). We did not observe genetic correlations of three COVID-19 traits with SLE significant in any specific functional annotation categories (Figure 2B).

**Figure 1.**
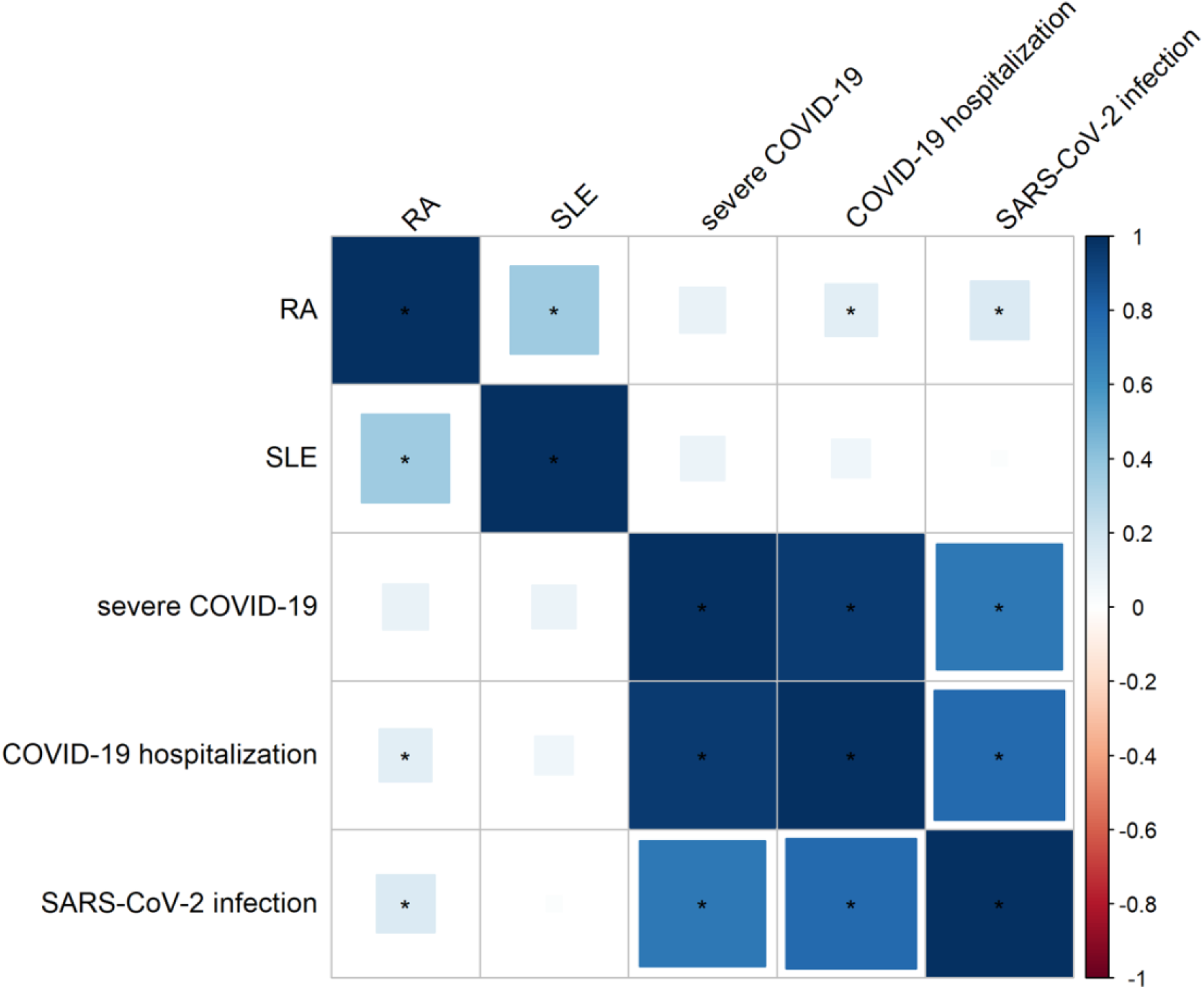
Genetic correlation of RA and SLE with COVID-19 related traits.

**Figure 2A.**
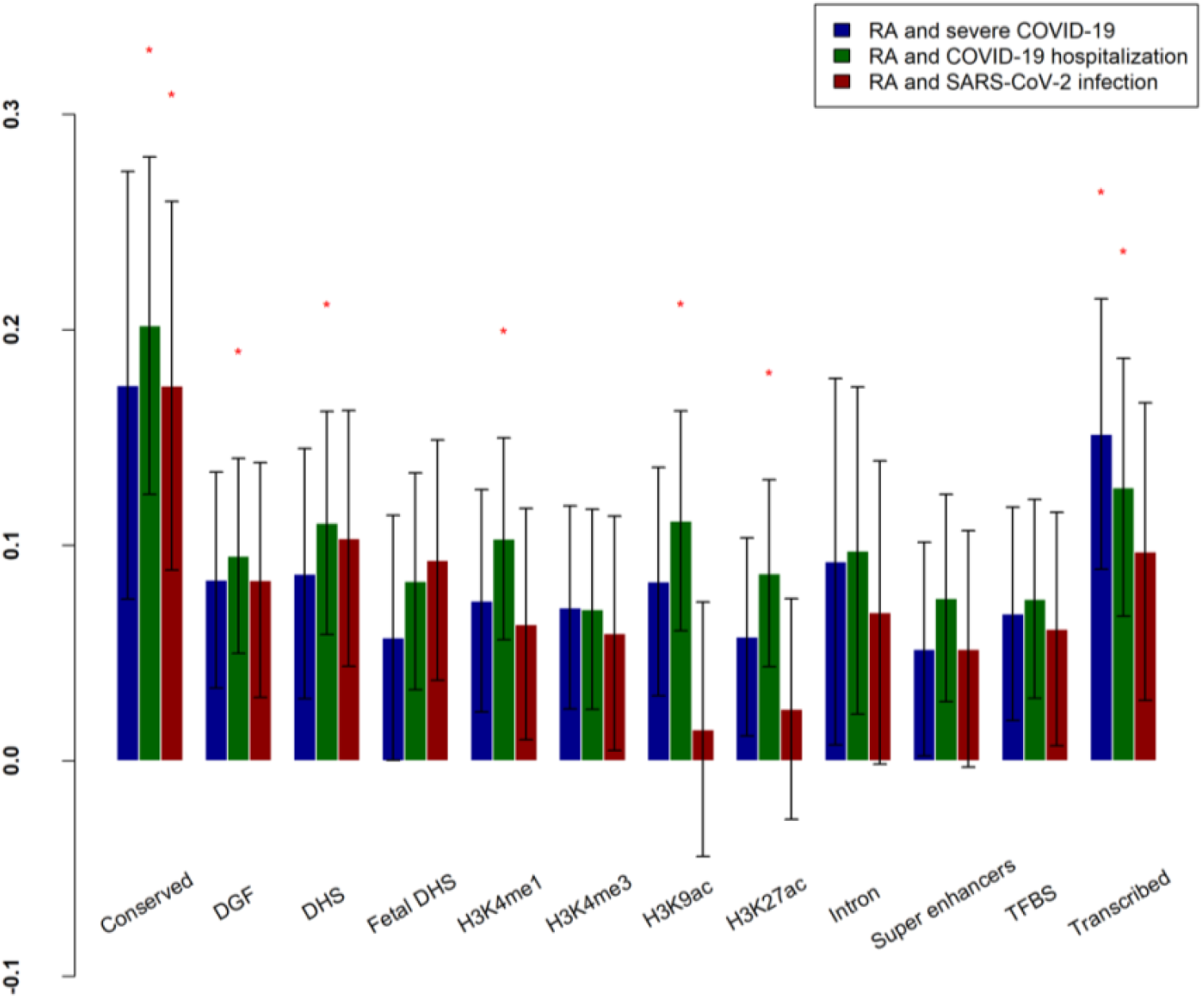
Partitioned genetic correlation between RA and COVID-19 related traits. The x-axis represents the 12 functional categories, and the y-axis represents the estimated partitioned genetic correlation. The significant functional categories (P<0.05) are starred. DGF: DNaseI digital genomic footprinting; DHS: DNase I hypersensitivity site; TFBS: transcription factor binding sites.

**Figure 2B.**
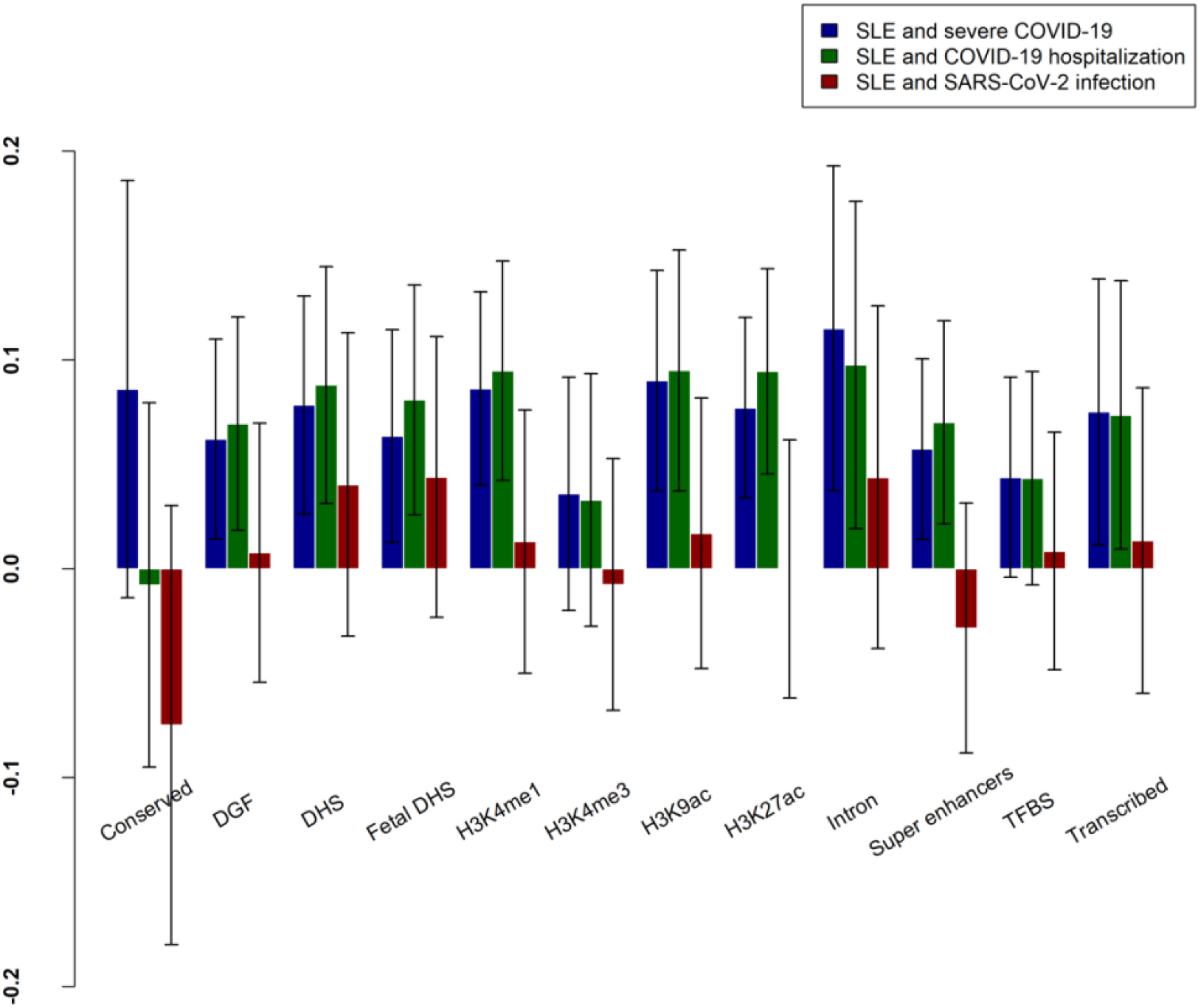
Partitioned genetic correlation between SLE and COVID-19 related traits. The x-axis represents the 12 functional categories, and the y-axis represents the estimated partitioned genetic correlation. The significant functional categories (P<0.05) are starred. DGF: DNaseI digital genomic footprinting; DHS: DNase I hypersensitivity site; TFBS: transcription factor binding sites.

### Multi-Trait Analysis of GWAS (MTAG)

Based on the updated COVID-19 host genetics initiative (HGI) release 7, there were 45 genome-wide significant (P<5×10^−8^) and uncorrelated (r^2^<0.01) loci for severe COVID-19, 46 for COVID-19 hospitalization and 26 for SARS-CoV-2 infection (Supplementary Table 3). We also extracted 102 and 68 genome-wide significant independent genetic loci for RA and SLE respectively using GWASs of European ancestry (Supplementary Table 4 and 5). Figure 3A and 3B displayed the Manhattan plots of all loci in the cross-trait meta-analysis. We did not identify additional genome-wide significant loci for COVID-19 traits using MTAG incorporating information from GWAS of RA or SLE.

**Figure 3A.**
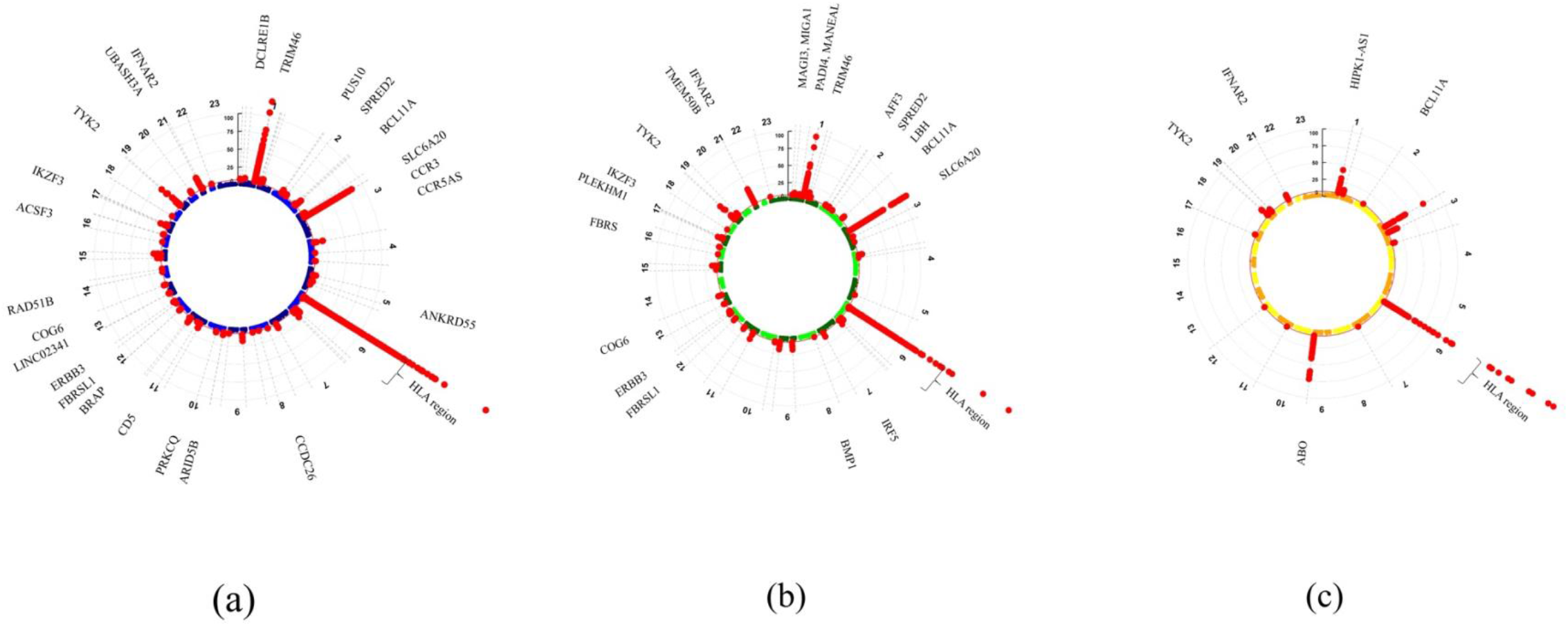
Circular Manhattan plot of cross-trait analysis for (a) RA and severe COVID-19, (b) RA and COVID-19 hospitalization, (c) RA and SARS-CoV-2 infection. Each point represents a SNP, and shared significant loci with meta-analysis P-value < 5×10^−8^ and single-trait P-value < 0.05 are colored in red. SNPs are arranged according to the chromosome position.

We identified 46 shared genetic loci associated with both severe COVID-19 and RA, 47 with COVID-19 hospitalization and RA, 19 with SARS-CoV-2 infection and RA, and 19 with severe COVID-19 with SLE, 24 with COVID-19 hospitalization and SLE, 11 with SARS-CoV-2 infection and SLE (P_meta_ < 5×10^−8^; single trait P <0.05) (Supplementary Tables 6-11).

We observe that the shared signals for all COVID-19 traits and RA were mainly mapped to the human leukocyte antigen (HLA) complex region at 6p22.1 to 6p21.3 (shown in Figure 3A and 3B), including class I region (*ZFP57, HCG4B, PSORS1C1, HLA-B, MICB*), class II region (*HLA-DQA1, HLA-DOB, HLA-DRB1, TAP2, PSMB9, COL11A2, TSBP1*), and class III region (*MSH5, EHMT2, C2, CFB, TNXB*). The strongest association signals for the three COVID-19 traits with RA were observed to be localized near the *HLA-DRB1* gene (index SNP: rs679242 for severe COVID-19, P_meta_=2.43×10^−345^; rs17425622 for COVID-19 hospitalization, P_meta_=5.44×10^−567^; rs2647062 for SARS-CoV-2 infection, P_meta_=1.49×10^−294^). For severe COVID-19, COVID-19 hospitalization and SARS-CoV2 infection with SLE, the strongest association signals were localized to the *TNXB* gene (index SNP: rs1150753, P_meta_=1.73×10^−69^), *CCR3* gene (index SNP: rs4443214, P_meta_=2.24×10^−57^) and *ABO* gene (index SNP: rs630014, P_meta_=4.10×10^−32^), respectively.

Notably, we identified 25 novel association genes for COVID-19 and RA which were not reported in previous COVID-19 GWAS, 22 of them were protein-coding genes, including *DCLRE1B, SPRED2, PUS10, ANKRD55, ARID5B, PRKCQ, CD5, ERBB3, BRAP, COG6, RAD51B, UBASH3A, MAGI3, MANEAL, MIGA1, PADI4, LBH, AFF3, IRF5, BMP1, FBRS, TMEM50B*. We identified 6 novel genes for COVID-19 and SLE, 4 of them were protein-coding genes, including *DCST1, KEAP1, ADAM15* and *SMG7*.

### Fine-mapping and co-localization analysis identify shared causal genetic variants

A list of SNPs in each shared locus from fine mapping was shown in Supplementary Table 12-17. Co-localization analysis shows that those colocalized genetic loci for RA and severe COVID-19 were rs2304256 (*TYK2*), rs12472359, rs9909593 (*IKZF3*), rs2855426 (*COL11A2*); for RA and COVID-19 hospitalization were rs2304256 (*TYK2*), rs3130564 (*PSORS1C1*), rs1265093 (*PSORS1C1*), rs9909593 (*IKZF3*), rs2306627 (*MANEAL*), rs7993214 (*COG6*); for RA and SARS-CoV-2 infection were rs2304256 (*TYK2*). And the colocalized loci for SLE and severe COVID-19 were rs16940677 (*CRHR1*); for SLE and COVID-19 hospitalization were rs62057151 (*CRHR1*), rs492602 (*FUT2*); for SLE and SARS-CoV-2 infection were rs2290859 (*NXPE3*), rs492602 (*FUT2*). (Table 1)

**Table 1.**
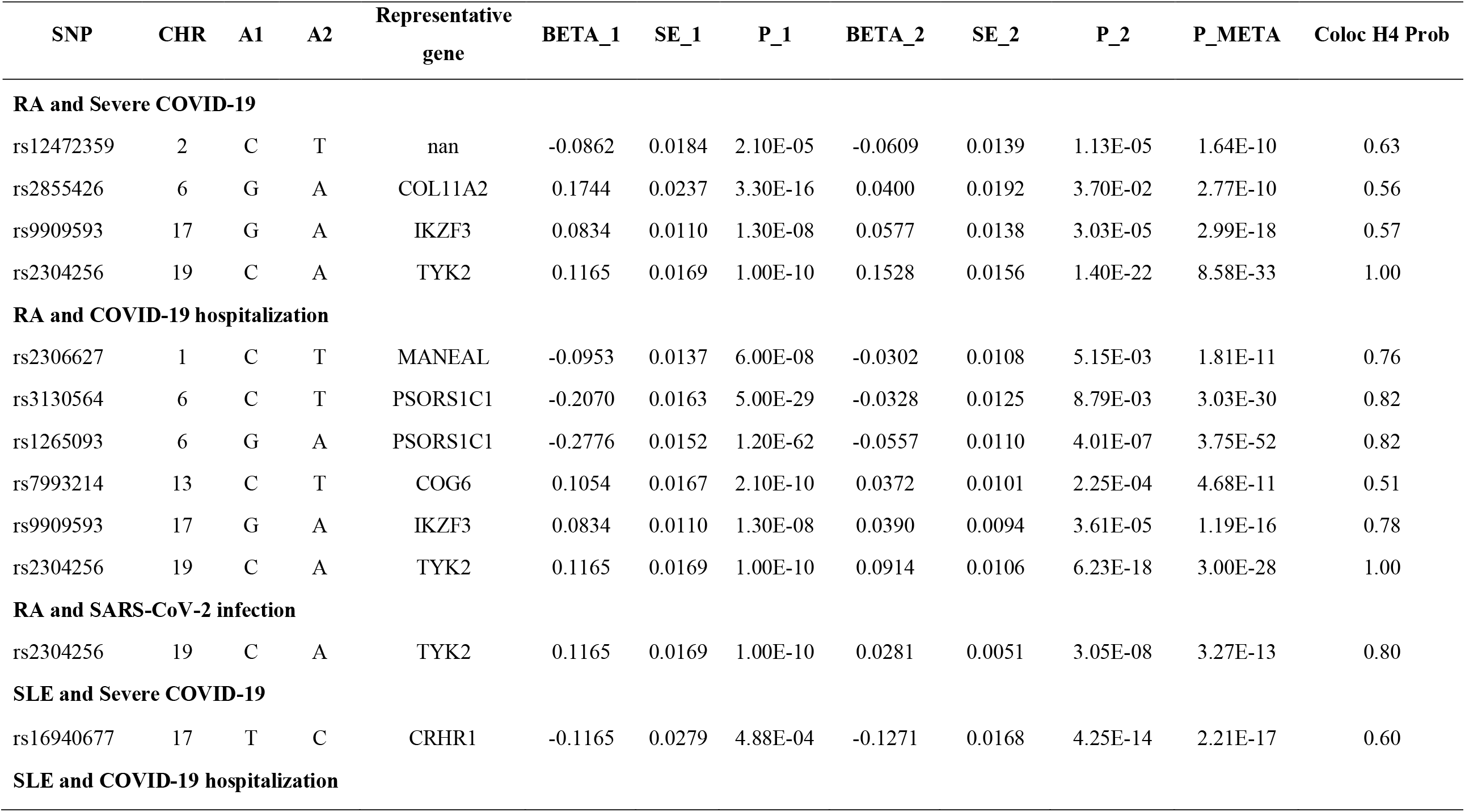
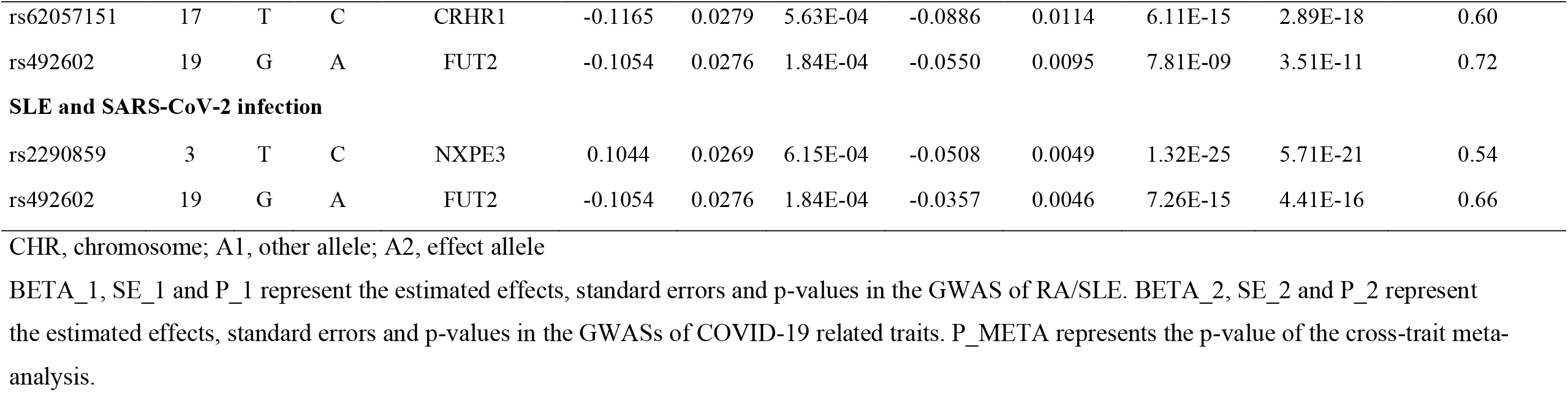
Co-localization analysis of COVID-19 and RA/SLE.

### GTEx tissue-specific expression analysis (TSEA) and over-representation enrichment analysis of shared genes

The identified shared genes for COVID-19 and RA are significantly enriched for expression in the spleen, lung, whole blood and small intestine; shared genes for COVID-19 and SLE are significantly enriched for expression in the spleen, lung and whole blood. (Figure 4A and 4B). Gene Ontology (GO) analysis results show that these shared genes were enriched in several immune response-related functional terms (Supplementary Table 18-23), such as “antigen processing and presentation”, “regulation of immune effector process”, “immune response-regulating signalling pathway”, “immunocyte (B cell, T cell, lymphocyte, leukocyte) mediated immunity”, and “response to chemokine/interferon”.

**Figure 4B.**
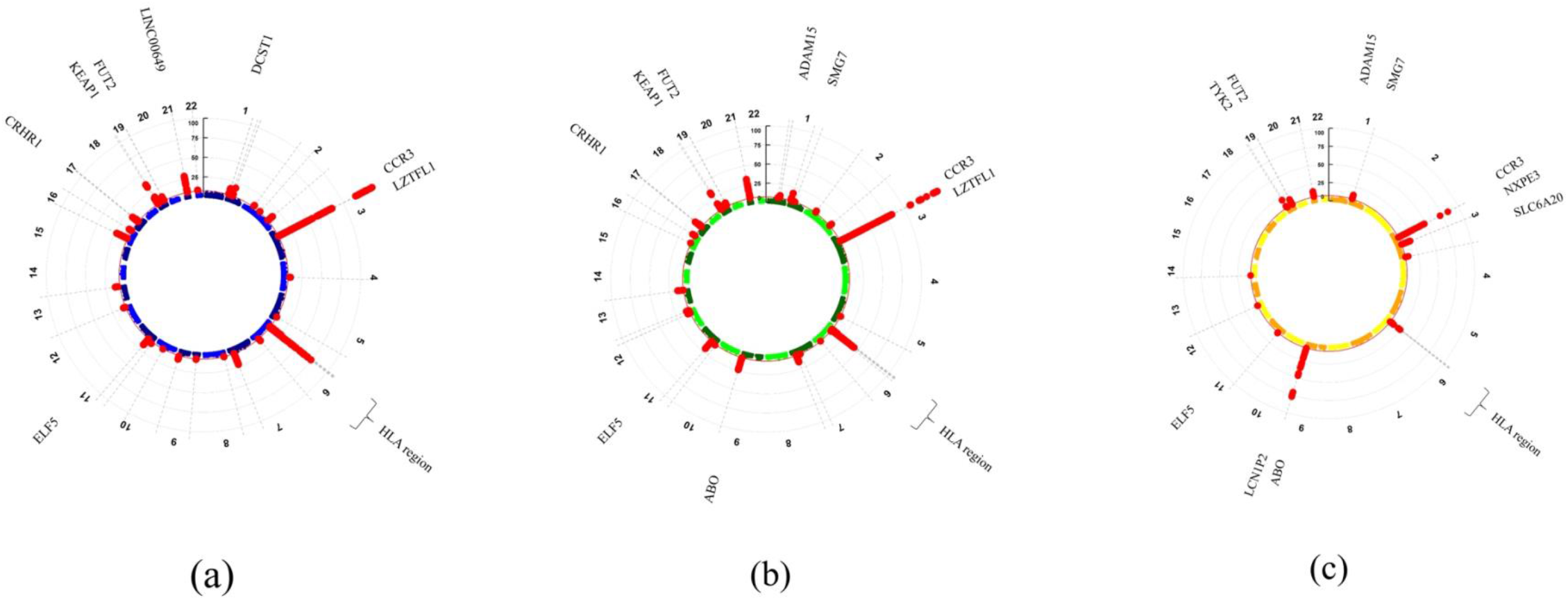
Circular Manhattan plot of cross-trait analysis for (a) SLE and severe COVID-19, (b) SLE and COVID-19 hospitalization, (c) SLE and SARS-CoV-2 infection. Each point represents a SNP, and shared significant loci with meta-analysis P-value < 5×10^−8^ and single-trait P-value < 0.05 are colored in red. SNPs are arranged according to the chromosome position.

### Mendelian randomization (MR) analysis

We used 36 SNPs for RA and 37 SNPs for SLE as instruments in the analysis on the association of RA/SLE with COVID-19 (Supplementary Table 24 and 25). We found that genetically predicted RA was positively associated with a higher risk of COVID-19 hospitalization (OR=1.05, 95%CI:1.02-1.09, P-value=1.53×10^−3^) (Figure 5A). In addition, the causal effect of RA on severe COVID-19 was significant when using the weighted mode method (OR=1.08, 95%CI: 1.01-1.16, P-value= 4.25×10^−2^), indicating a positive association of RA on severe COVID-19 (Supplementary Figure 1A). We found null associations of genetically predicted SLE with COVID-19 (Figure 5B). In the other direction, using 26 SNPs as instruments for severe COVID-19, 27 SNPs for COVID-19 hospitalization and 11 SNPs for SARS-CoV-2 infection (Supplementary Table 26-28), we found null associations of genetically predicted COVID-19 with RA or SLE (Figure 6A and 6B), and the results are robust to different MR methods (Supplementary Figure 2A and 2B).

**Figure 5A.**
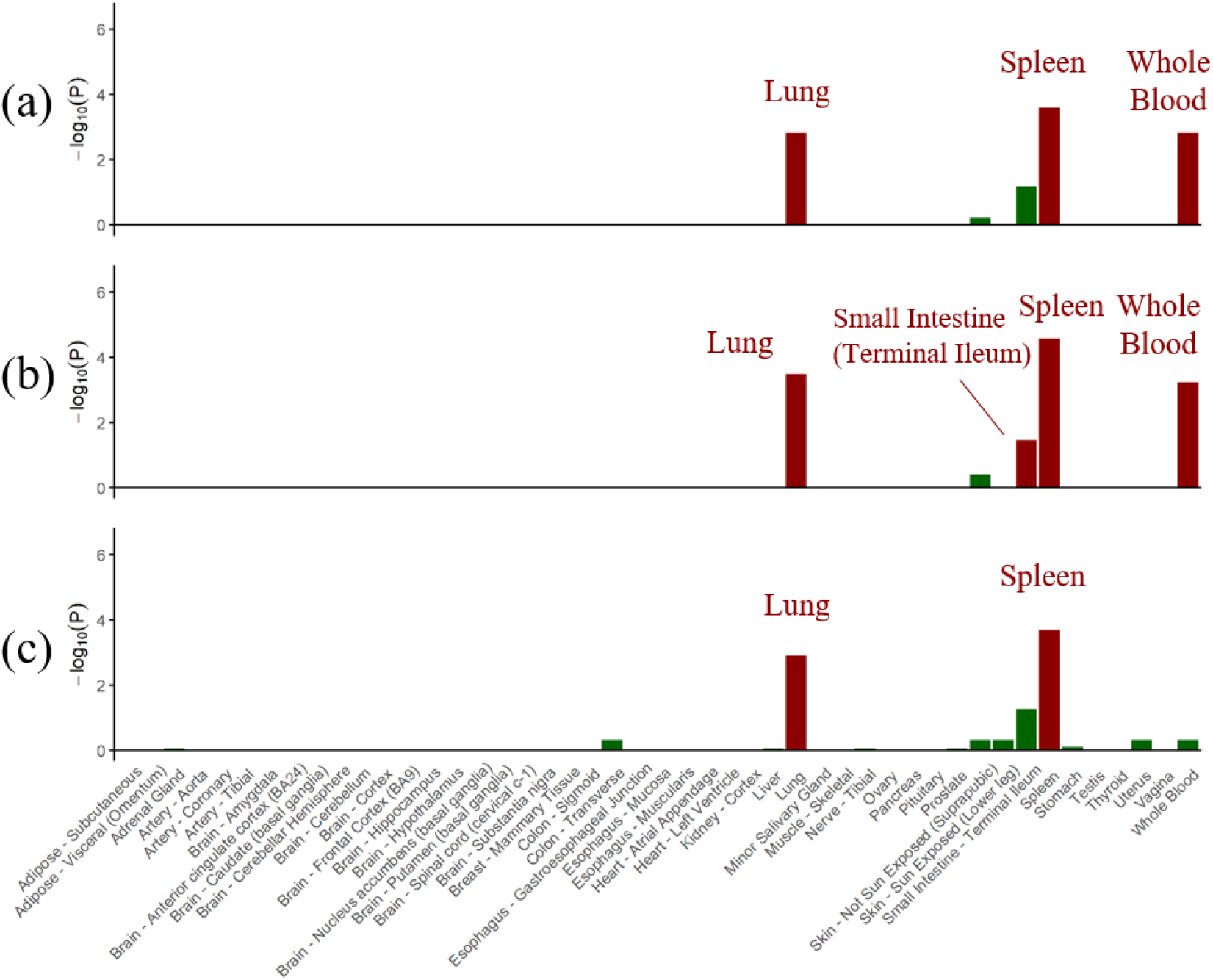
GTEx tissue enrichment analysis for expression of shared significant genes (meta-analysis P-value < 5×10^−6^) for (a) RA and severe COVID-19, (b) RA and COVID-19 hospitalization, (c) RA and SARS-CoV-2 infection. P-values of Fisher’s exact test after Benjamin-Hochberg correction are presented in –log_10_ scale. Red represents significant tissue enrichment.

**Figure 5B.**
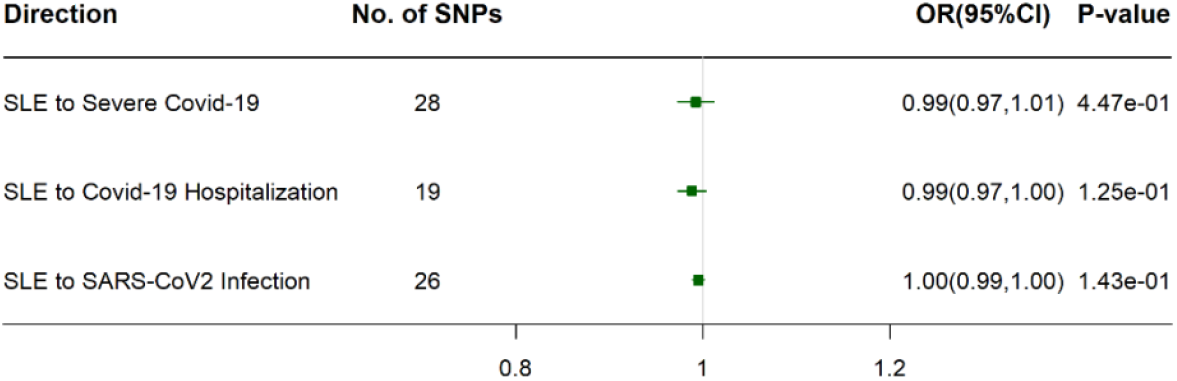
MR analysis of SLE to COVID-19 related traits using IVW method. Causal effect estimates are presented as odds ratios (OR) with 95% confidence intervals (CI).

**Figure 6A.**
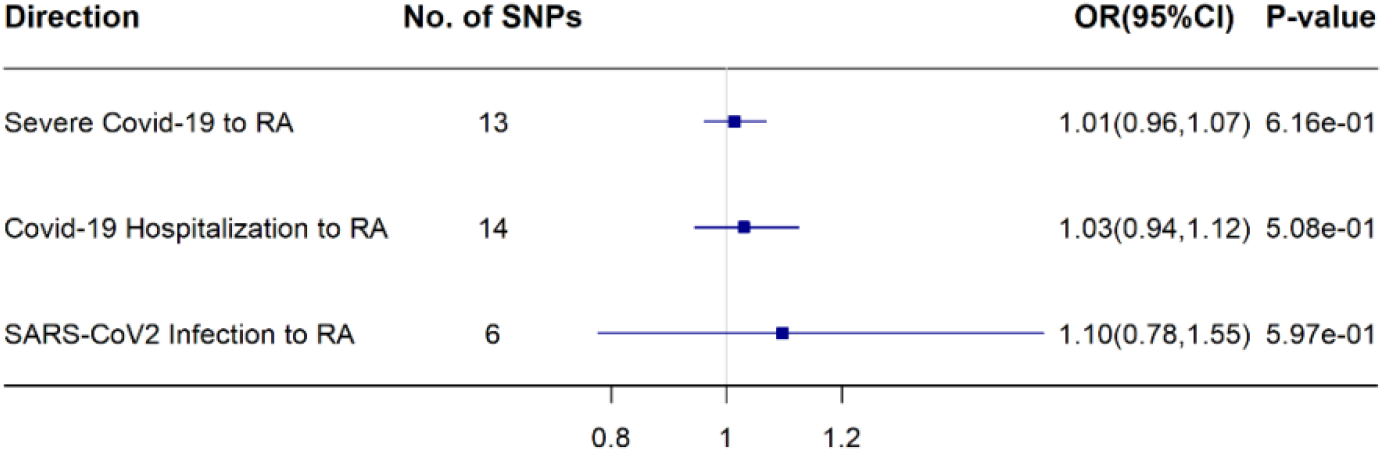
MR analysis of COVID-19 related traits to RA using IVW method. Causal effect estimates are presented as odds ratios (OR) with 95% confidence intervals (CI).

**Figure 6B.**
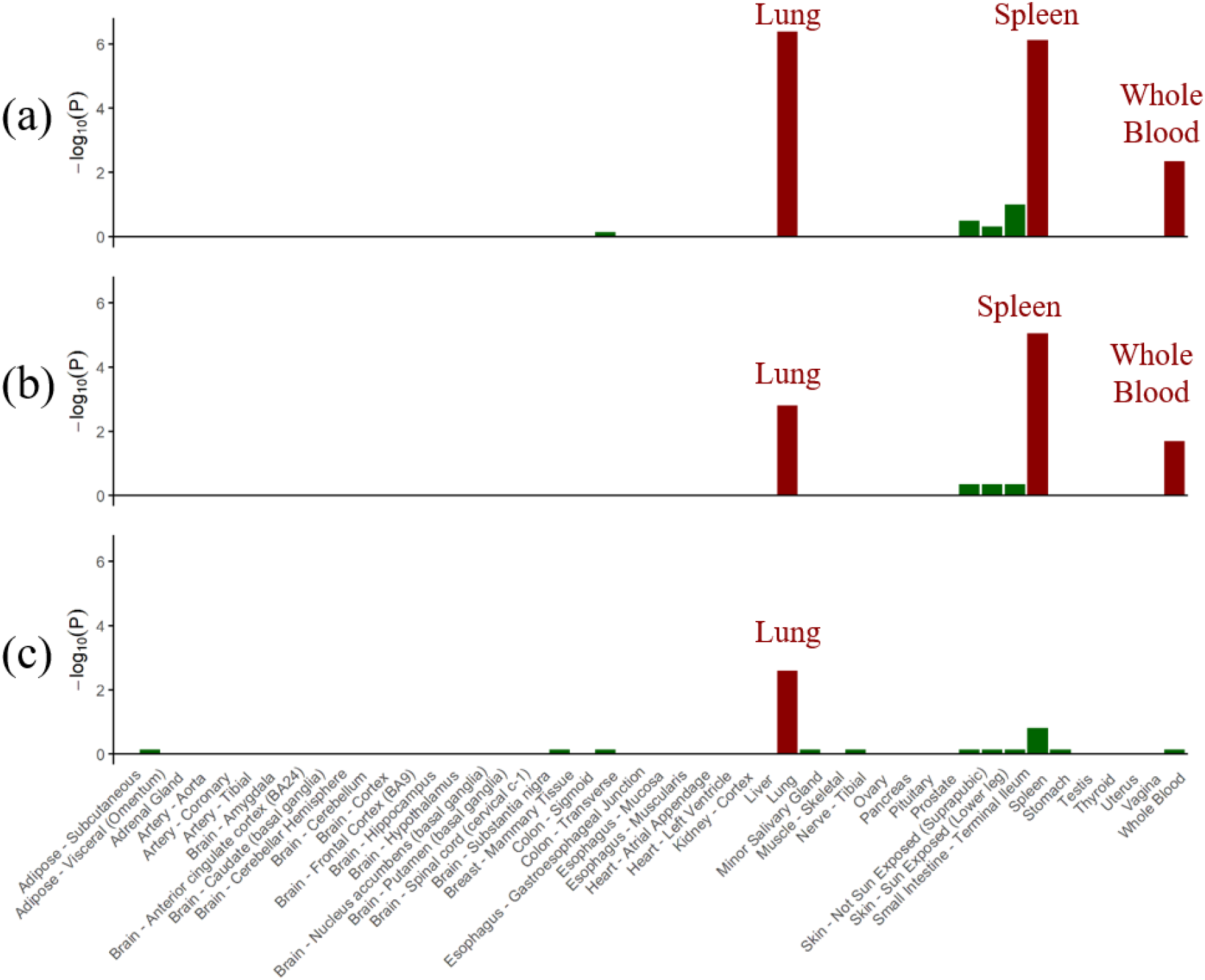
GTEx tissue enrichment analysis for expression of shared significant genes (meta-analysis P-value < 5×10^−6^) for (a) SLE and severe COVID-19, (b) SLE and COVID-19 hospitalization, (c) SLE and SARS-CoV-2 infection. P-values of Fisher’s exact test after Benjamin-Hochberg correction are presented in –log_10_ scale. Red represents significant tissue enrichment.

## Discussion

In this study, we disentangled the shared genetic etiology and causality of three COVID-19 traits with RA and SLE using large-scale GWAS data from the most recent COVID-19 HGI and large consortia on RA and SLE. For the first time, we found positive genetic correlations and further identified shared genetic loci of three COVID-19 traits (severe COVID-19, COVID-19 hospitalization and SARS-CoV-2 infection) with RA and SLE. The shared genetic loci were significantly enriched in several tissues, including spleen, lung, whole blood and small intestine. We also applied bi-directional MR analysis to examine the causal relationship, and found that RA may increase the risk of severe COVID-19 and COVID-19 hospitalization.

Our LDSC analysis showed a significant positive genetic correlation of COVID-19 hospitalization and SARS-CoV-2 infection with RA, and partitioned LDSC analysis found the genetic correlation of severe COVID-19 with RA were positively significant in the transcribed region. Although the positive genetic correlations of SLE with COVID-19 traits were not significant at the 0.05 level, we identified shared loci in cross-trait meta-analysis, supporting that they may have shared genetic basis.

In the cross-trait meta-analysis, we found that most of the genetic loci shared by RA and SLE with COVID-19 related traits were located in the *HLA* region, which strengthened the evidence for a role of *HLA* region in COVID-19 and autoimmune diseases In particular, we observed the strongest shared signals for three COVID-19 traits with RA are located near the *HLA-DRB1* gene region, encoding HLA class II molecules, which are associated with antigen presentation and multiplication of T-helper cells, and is important in initiating immune responses.^10,11^ *HLA* region has been known to be related to the regulation of immune function, and HLA-II alleles have been reported to be associated with autoimmune diseases, including RA and SLE.^12,13^ *HLA* complex has also been reported to be associated with COVID-19 susceptibility and severity,^14-17^ primarily by influencing the binding affinity to viral peptides on the surface of SARS-CoV-2.^14,18^ These evidence suggest that dysregulated immune-inflammatory responses play an important role in the pathogenic mechanisms of COVID-19, RA and SLE. This is consistent with clinical findings that hyperactivation of immune responses is the principal feature of COIVID-19 and the cytokine hyperactivation in COVID-19 is similar to that in RA.^19^

Some other identified shared genes were also involved in the regulation of immune function. For example, among the genes shared by COVID-19 with RA, *IKZF3* and *BCL11A* genes are known to be involved in B- and T-lymphopoiesis and differentiation, which are essential for the establishment of adaptive immune system;^20,21^ *TYK2* and *IFNAR2* genes mediate the JAK-STAT signaling pathway, regulating interferon and cytokine related responses.^22,23^ Notably, the shared signals of all three COVID-19 traits with RA and SLE included the 3p21.31 gene region (containing *SLC6A20, LZTFL1* and *CCR3* genes). This region has been reported to be associated with severe COVID-19 in the first COVID-19 GWAS, and some putative coreceptors genes (*LZTFL1, SCL6A20*) and chemokines and their receptors genes have been mentioned.^24^ *CCR3*, the gene with the strongest shared signals for COVID-19 hospitalization and SLE, encodes the receptor for CC-type chemokines that are involved in the accumulation and activation of inflammatory cells such as eosinophils.^25^

Interestingly, among the genes shared by COVID-19 and SLE, we identified some genes related to haemocyte function. We found strong shared signals for SARS-CoV-2 infection and COVID-19 hospitalization with SLE in the *ABO* gene and *FUT2* gene. Previous GWAS have shown that *ABO* was a locus associated with COVID-19 susceptibility and severity,^26^ mechanisms supporting these findings may be related to the ABO group per se (e.g. blood group polymorphisms leading to differential viral adherence or interaction with ACE2) and associations with coagulation factors.^27,28^ *FUT2* is a fucosyltransferase gene involved in ABO blood group antigen synthesis, and has been found to be associated with severe COVID-19 and SARS-CoV-2 infection.^15^ We also found a novel shared gene, *ADAM15*, whose expression serves as an adhesion receptor for platelets, plays a role in platelet activation and thrombus formation.^29^ Hematological alterations are also common in patients with SLE,^30,31^ these evidence suggests the important function of erythrocyte surface antigens and coagulation in COVID-19 and SLE co-morbidities.

In addition to immune and coagulation functions, we found some genes shared between severe COVID-19, COVID-19 hospitalization and RA are associated with bone metabolism, such as genes at 1p13.2 (*DCLRE1B* and *MAGI3*), *LBH* gene and *BMP1* gene. Locus 1p13.2 is an important region to regulate RA, and *MAGI3* were found to be associated with osteoblast proliferation, differentiation and bone formation.^32^ *BMP1* (Bone Morphogenetic Protein 1) encodes a protein that is capable of inducing formation of cartilage in vivo,^33^ and *LBH* expression was found to influence bone formation.^34^ Consistently, osteoporosis and osteonecrosis are common sequalae in severe COVID-19 patients.^35^ Furthermore, we also found shared signals for severe COVID-19 and RA in genes associated with intestinal diseases, including *PUS10, PRKCQ* and *ANKRD55*. They are identified to be risk genes for intestinal inflammatory diseases such as Crohn’s disease, celiac disease, and ulcerative colitis.^36-38^ Symptoms of gastrointestinal inflammation or injury are frequently observed in patients with COVID-19 and RA.^39^

Furthermore, by means of colocalization analyses, we identified likely causal variants shared by COVID-19 with RA or SLE, were localized to *TYK2, IKZF3, COL11A2, PSORS1C1, MANEAL, COG6* genes for COVID-19 with RA, and *CRHR1, FUT2, NXPE3* genes for COVID-19 with SLE. As we discussed previously, some of these genes are involved in immune function and inflammation (such as *TYK2*) or coagulation (such as *FUT2*). This is also in line with the findings from enrichment analysis. Our TSEA reported that shared genes for three COVID-19 traits with RA/SLE were significantly enriched for expression in the spleen, whole blood, lung and small intestine, which are important for immune function, inflammation and coagulation functions, supporting that COVID-19 and RA/SLE have shared genetic etiology in these aspects. Importantly, these results also provide more insights into disease pathogenesis and pinpoint targets for therapeutic development or drug repurposing. For example, the pan-JAK inhibitor drug Baricitinib used for RA and atopic dermatitis,^40^ which can inhibits TYK2, has shown promising results in trials for its efficiency in COVID-19.^22,41^ The expression of *PSORS1C1* gene regulate the secretion of IL-17,^42^ and IL-17 inhibitors for the treatment of psoriasis and RA are considered to be a promising therapeutic option to terminate COVID-19.^43^ *CRHR1*, gene of receptor binding, is associated with the efficacy of corticosteroids.^44^ Consistently, an open-label randomized controlled trial showed that taking dexamethasone, a corticosteroid, may lower the risk of 28-day mortality by 17% COVID-19-hospitalized patients.^45^

Our bi-directional Mendelian randomization study adds to the limited evidence on the causal relationship between RA and COVID-19. For the first time, we showed that genetically predicted RA is associated with higher risk of COVID-19 hospitalization and possibly with severe COVID-19. The positive association is consistent with the findings from large cohort studies showing patients with RA are at higher risk of hospitalization and mortality of COVID-19. Currently, the Advisory Committee on immunization have not put people with autoimmune diseases at high priority (https://www.cdc.gov/mmwr/volumes/69/wr/mm695152e2.htm?s_cid=mm695152e2_w), possibly due to the lack of solid evidence. Our findings suggest that patients with RA may be promoted to take vaccines, given the potential higher risk of COVID-19 hospitalization and severe COVID-19. Our bi-directional MR does not suggest a higher risk of RA or SLE associated with COVID-19, which do not support the concern that COVID-19 may trigger autoimmunity and thereby induce RA or SLE. Partly consistent with our findings, a recent study shows that taking SARS-CoV-2 mRNA vaccines lead to development of antibodies in immunosuppressed patients without induction of disease flares.^46^

Our study is the first to explore the common genetic architecture between RA or SLE with COVID-19-related traits. We used data from the COVID-19 HGI release 7 summary statistics, the largest and latest GWAS of COVID-19 available, and well-powered meta-analysis of multiple GWAS studies, allowing for reliability and genetic robustness of the study. Our study provides timely evidence directly related to the genetic etiology of these diseases. We acknowledge a few limitations of our study. First, we used RA, SLE and COVID-19 GWAS data of European ancestry, the generalizability of our results should be considered with caution. Second, only summary statistics for COVID-19 are available, so we could not test whether the shared genetic architecture of COVID-19 with RA and SLE may vary by sex or age. Third, the marginal, positive association of RA with severe COVID-19 might have occurred by chance, or might be due to a lack of power. So, replication in a larger GWAS of severe COVID-19 will be needed. Finally, although our study provides evidence of common genetic basis between COVID-19 and RA/SLE, the underlying mechanism remains unclear. Further biological experiments are warranted.

In summary, our findings provide strong evidence of genetic correlations and causal association between COVID-19 and RA as well as SLE. We identified their shared genetic loci, with mapped genes related to immunity, inflammation and coagulation function. This work reinforces the idea that COVID-19 and RA/SLE share common biological processes and may help to pinpoint therapeutic targets. Our study also indicated that RA might be linked to higher risk of severe COVID-19 and COVID-19 hospitalization, emphasizing the importance of preventing severe COVID-19 in people with RA.

## Methods

### Study population

Our study is based on the summary level data from well-powered meta-analysis of multiple GWAS studies. Genetic associations of COVID-19 were obtained from the latest freeze 7 data (release date: April 08, 2022) of the COVID-19 host genetics consortium (https://www.covid19hg.org/results/). We studied three related traits of COVID-19: (1) Severe COVID-19, defined as COVID-19-confirmed individuals with very severe respiratory symptoms or those who died from the disease (up to 13,769 cases and 1,072,442 controls).

(2) COVID-19 hospitalization, defined as individuals who were hospitalized for related infection symptoms (up to 32,519 cases and 2,062,805 controls). (3) SARS-CoV2 infection, defined as all individuals who reported positive for SARS-CoV2 infection regardless of symptoms and hospitalization (up to 122,616 cases and 2,475,240 controls).

Genetic associations with risk for RA were measured on 14,361 cases and 43,923 controls of European ancestry and obtained from a GWAS meta-analysis with 18 studies included,^47^ and for SLE were measured on 7,219 cases and 15,991 controls of European ancestry and obtained from the University of Michigan Health and Retirement Study (HRS).^48^

### Linkage disequilibrium score regression (LDSC)

We applied LDSC to calculate the heritability for a single trait and genetic correlation between two traits using the GWAS summary statistics and LD scores of 1000G European ancestry reference.^49^ By running a regression of the test statistics on LD scores, LDSC provides an estimate of the heritability or genetic correlation. LDSC can provide accurate estimates even if the test statistics are inflated due to polygenicity.^15^

### Partitioned LDSC analysis

We performed partitioned LDSC analysis to estimate the genetic correlation between two traits within each of the following 12 functional categories^50^: conserved region, DNaseI digital genomic footprinting region (DGF), DNase I hypersensitivity sites (DHS), fetal DHS, H3K4me1, H3K4me3, H3K9ac and H3K27ac, intron region, super enhancers, transcription factor-binding site (TFBS) and transcribed region. With the partitioned genetic correlations, we can find out which functional categories account for the majority of the overall genetic correlation.

### Multi-Trait Analysis of GWAS (MTAG)

We conducted a meta-analysis with multiple GWASs to identify novel SNPs with strong signals using the Multi-Trait Analysis of GWAS (MTAG).^51,52^ MTAG re-estimates the effect sizes for each COVID-19 trait by incorporating the weighted sum of GWAS estimates for RA/SLE. We also used MTAG to conduct cross-trait meta-analysis to detect the loci that are significant in both traits, which utilizes sample size-weighted, fixed-effect model together with genetic covariance modeling from all sources to combine evidence of the association between individual variants for RA/SLE and COVID-19.

### Fine-mapping and co-localization analysis

After identifying the significant shared loci between two traits, we applied one Bayesian algorithm, namely Probabilistic Identification of Causal SNPs (PICS), to find out a 99%-credible set of SNPs for each independent shared genetic variant within 500 kb of the index SNP.^53^ Then we conducted co-localization analysis to check whether COVID-19 and RA/SLE association signals co-localized at shared loci, by calculating the posterior probability that two traits shared the same causal variant (P(H4)).^54^ If P(H4)>0.5 for any SNP, we labelled it as a co-localized genetic variant.

### Tissue-specific expression analysis (TSEA) and over-representation enrichment analysis

We also performed TSEA with the HUGO Gene Nomenclature Committee (HGNC) name of genes that correspond with the shared loci in cross-trait meta-analysis. TSEA aims to test whether the shared genes are enriched in a specific tissue and often requires external reference panels. The R package ‘deTS’ uses GTEx RNA-seq data and ENCODE panel as the reference panel, and the corresponding z-score is calculated for each tissue.^55^ Furthermore, we used the same gene list to analyze the enrichment in Gene Ontology (GO) biological process by the WEB-based GEne SeT AnaLysis Toolkit.^56^

### Mendelian randomization (MR) analysis

We conducted a bidirectional MR analysis to assess the causal relationship between two traits. For IV selection, we first excluded SNPs in *HLA* gene region, *ABO* gene and *FUT2* gene because genetic variants in these regions might be susceptible to horizontal pleiotropy. Then we extracted genome-wide significant (P < 5 × 10^−8^) and independent (r^2^ < 1 × 10^−4^) SNPs as candidate IVs. For each exposure-outcome pair, we applied two-stage hard thresholding with voting to select valid IVs for MR analysis.^57^ Our main results were based on the inverse variance weighted (IVW) method,^58^ and sensitivity analyses were performed using the following methods: weighted median,^59^ weighted mode,^60^ MR-Egger,^61^ and MR-RAPS ^59,61-63^.

The details of those analysis methods were provided in the Supplemental Note.

## Data Availability

Genetic associations with severe COVID-19, COVID-19 hospitalization and SARS-CoV-2 infection were obtained from COVID-19 host genetics consortium GWAS meta-analyses round 7, downloaded from https://www.covid19hg.org/results/r7/. Genetic associations with RA and SLE were obtained from international consortia and can be downloaded from http://insidegen.com/ and http://plaza.umin.ac.jp/~yokada/datasource/software.htm.

http://insidegen.com/

http://plaza.umin.ac.jp/~yokada/datasource/software.htm

## Statements and Declarations

### Author Contributions

The manuscript was drafted by XH, with great help of JVZ and ZL. Data analysis was conducted by MY and YG. XH and MY interpreted the results, with the help of ZL and JVZ. ZL and JVZ developed the study conception, designed the analysis, and critically revised the manuscript for important intellectual content. All authors read and approved the final manuscript.

### Ethics approval

The study is an analysis using publicly available summary data that does not require ethical approval.

## Acknowledgement

Genetic associations with COVID-19, RA and SLE were obtained from the COVID-19 host genetics initiative and international consortia. The authors would like to thank all participants in the study and investigators for sharing the valuable data.

---

**Figure 5A.**
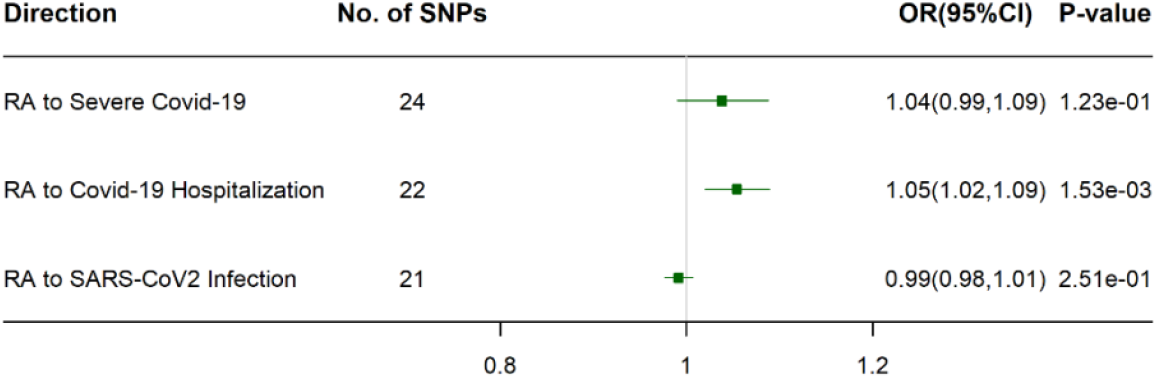
MR analysis of RA to COVID-19 related traits using IVW method. Causal effect estimates are presented as odds ratios (OR) with 95% confidence intervals (CI).

**Figure 6B.**
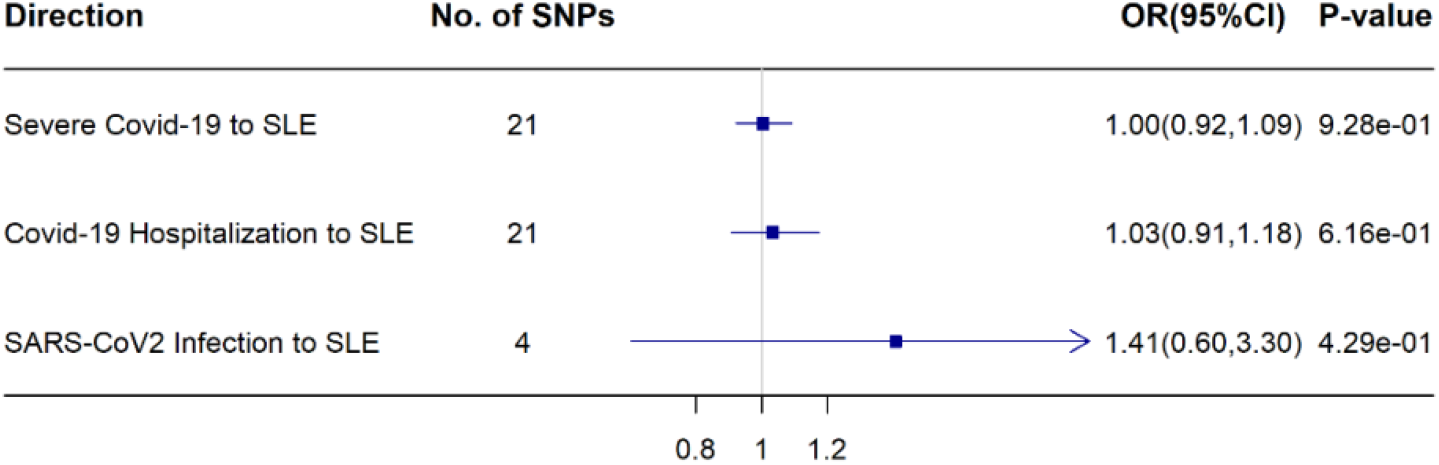
MR analysis of COVID-19 related traits to SLE using IVW method. Causal effect estimates are presented as odds ratios (OR) with 95% confidence intervals (CI).

